# Adherence, Adverse Events and Viral Control among Children and Adolescents with HIV in Zambia Switched to an Integrase Inhibitor Regimen

**DOI:** 10.1101/2024.09.17.24313837

**Authors:** David R. Bearden, Kristen Sportiello, Milimo Mweemba, Frank Lungu, Sylvia Mwanza-Kabaghe, Gretchen Birbeck

## Abstract

**Background:** Based on recent World Health Organization recommendations, there has been a large-scale transition in Sub-Saharan Africa to integrase inhibitor (II)-based antiretroviral therapy (ART) regimens.

**Setting:** This study was conducted at an urban referral center in Lusaka, Zambia.

**Methods:** This study included 297 children and adolescents with HIV (CAWH) on ART for one year prior to enrollment and followed for 1-4 years after enrollment. ART adherence, ART regimen, and viral load were assessed periodically. Structured interviews were conducted with a subset of 95 children to assess adherence barriers and side effects.

**Results:** Children on protease inhibitor (PI)-based regimens were more likely to report adherence problems than children taking II- or Efavirenz-based regimens (10% vs. 28%, p=0.03) and noted more days with missed doses (median 1 vs. 0, p=0.02). In interviews, the most common reasons given for poor adherence included bad medication taste, not being home when medications were due, and perceived side effects. The PI group was more likely to report that taste was a problem affecting adherence (22% vs. 4%, p=0.05) and headache as an ART side effect (17% vs. 4%, p=0.05). Switching from a PI- to an II-based regimen was associated with improved adherence (72% vs. 92%, p=0.01) and an undetectable viral load (67% vs. 78%).

**Conclusions:** Switching CAWH from PI-based to II-based regimens has many advantages including superior side effect profiles, adherence, and viral suppression. PI taste aversion may be a significant contributor to pediatric adherence issues. Palatability should be considered in pediatric HIV drug development.

## Introduction

There are approximately 38 million individuals living with human immunodeficiency virus (HIV) with two thirds of those living in Sub-Saharan Africa (SSA) [32, 42-43]. Despite the widespread use of antiretroviral therapy (ART), HIV remains one of the leading causes of death in low- and middle-income countries [33]. There are approximately 66,000 adolescents living with HIV in Zambia, although currently only 60% are estimated to be on ART [34]. High rates of ART adherence are necessary to prevent recurrent viremia and the development of resistance mutations [9-13]. ART adherence is particularly important in SSA given relatively few choices for salvage regimens should virologic failure occur [14-16].

The HIV-associated Neurocognitive Disorders in Zambia (HANDZ) study is an ongoing prospective longitudinal cohort study following 300 children and adolescents with HIV (CAWH) with initial recruitment taking place from 2017-2018. At that time, the majority of CAWH in Zambia were taking an ART regimen that included either Efavirenz (EFV) or a boosted protease inhibitor (PI) along with a nucleotide reverse transcriptase inhibitor (NRTI) backbone. In 2019, the World Health Organization (WHO) issued revised guidelines recommending that the integrase inhibitor (II) Dolutegravir should be preferentially utilized in children over EFV or PIs (along with an optimized NRTI backbone) due to lower costs, superior efficacy, availability of once-daily fixed-dose combinations, and a potentially better long-term side effect profile [1-2]. Furthermore, the WHO recommended that countries consider switching children who were stably virally suppressed on their current regimens to a dolutegravir-based regimen [2]. The WHO did acknowledge the risks of this approach, including the possibility of children developing new side effects and discontinuing treatment or reducing adherence [2]. Several studies in adults (21-25) had previously demonstrated increased adverse events and discontinuation due to side effects, particularly neuropsychiatric side effects or skin rashes, when switching from a PI- or NNRTI-based regimen to an II-based regimen. However, there are very limited data on the outcomes of this switch in children.

In Zambia, most children followed in the Pediatric Center of Excellence (PCOE) in the capital city of Lusaka were switched to dolutegravir following the WHO recommendations between 2019 and 2023. In the current study, we sought to investigate the effects of this switch on viral suppression, adherence, and side effect burden among CAWH followed in the HANDZ study. Prior HANDZ work identified lower rates of adherence and higher rates of reported side effects with PI-based regimens compared to others (unpublished data). We hypothesized that switching from a PI-based regimen to a regimen based on the II dolutegravir would be associated with improved adherence and reduced reported side effects.

## Methods

### Study design and setting

This study was conducted as a substudy of the HIV-associated Neurocognitive Disorders in Zambia (HANDZ) study. Details of the HANDZ study have previously been published (33-39). Briefly, the HANDZ study is an ongoing prospective longitudinal cohort study taking place in Lusaka, Zambia with 300 children and adolescents living with HIV and 300 HIV-uninfected controls enrolled. The first wave of the HANDZ study recruited 208 participants with HIV aged 8-18 years from 2017-2018. The second wave of the HANDZ study recruited an additional 92 participants with HIV (aged 3-18 years) from 2019-2022. As part of the HANDZ study, children are seen every 3 months for standardized interviews, adherence assessments and neuropsychological testing. The Collaborative HIV Investigations of Antiretroviral Neuropsychiatric Toxicities in Zambia (CHANTZ) study is a mixed methods substudy nested within the HANDZ study focused on neuropsychiatric side effects of ART and includes 95 total participants, all with perinatally acquired HIV. The CHANTZ study conducted detailed interviews regarding barriers to adherence and ART side effects. In the present analysis, we evaluated 3 timepoints: a baseline visit (timepoint 1), a second visit taking place 2 years after the baseline visit at which point participants were recruited into the CHANTZ substudy (timepoint 2), and a third visit completed approximately 1 year later (timepoint 3).

### Inclusion/Exclusion Criteria

To be enrolled in the HANDZ study, participants with HIV were required to have received ART for at least one year prior to enrollment and to have at least one living parent available for consent. Participants with a history of central nervous system opportunistic infections or chronic medical or psychiatric conditions that could preclude participation in the study were excluded. For the CHANTZ substudy reported here, enrollment was sought from all HANDZ participants taking PIs as well as age-matched HANDZ participants taking ART with no PI exposure in the previous five years. Written informed consent was required from the parent/guardian for inclusion in the study and, where appropriate, participant assent was also necessary.

### Data Collected

Participant and parent interviews as well as chart reviews of electronic and paper medical records were utilized to collect data on demographic and medical history. Adherence was assessed using standardized questionnaires adapted from the Pediatric HIV AIDS Cohort Study (PHACS) in addition to pill counts and viral loads. Data regarding side effects and barriers to adherence were collected using structured questionnaires developed for the study and administered to each child or adolescent as well as the parent unless the child or adolescent attended clinic visits independently without a parent. Interviews were conducted by local Zambian staff fluent in English and the appropriate local languages.

### HIV Data

HIV history was retrieved through participant and parent interviews alongside chart reviews. Measures of current and historical HIV disease severity included current and lowest recorded CD4 counts and percentages, viral loads prior to enrollment, current and worst recorded WHO Stage, previous hospitalizations, and history of opportunistic infections. Current and prior ART was verified through interviews and chart review. CD4 counts and viral loads were measured annually, with viral loads <50 copies considered undetectable.

### Sample Size

Sample sizes for the parent HANDZ study were based on simulations for each of the aims of the study with the goal of ensuring model stability and avoiding overfitting regression models. The sample sizes for CHANTZ were determined by recruiting all available participants in HANDZ taking PIs at baseline and an equal number of controls taking ART that did not include a PI at baseline with no PI exposure in the five years before baseline.

### Data Analysis

Statistical analyses were performed using Stata 16 (Stata Corp LP, College Station, TX). Student’s t-test or a chi-square test was used to compare each baseline demographic or clinical characteristic by ART regimen (efavirenz (EFV) vs. PI vs. II). For each outcome of interest, a bivariate analysis of association between the exposure (i.e., treatment with Efavirenz, PI, or II) and the outcome was completed using a Pearson’s chi-square test for dichotomous variables and a one-way ANOVA or Students t-test for continuous outcome measures. Baseline adherence variables were evaluated between groups using linear regression for continuous variables and logistic regression for dichotomous or categorical variables, with age, HIV-related disease severity at baseline, and prior ART failure treated as potential confounders. The effect of switching regimens was evaluated using linear mixed effects models, controlling for potential confounders. Directed Acyclic Graphs (DAGs) were used to visualize the causal model using Daggity (www.daggity.net, 44-46), and the minimal sufficient adjustment set suggested by the DAG was used to select variables to include in the model (see Figure 1). HIV disease severity, prior ART failure, age, and baseline adherence were included as possible confounders. In addition, structured qualitative interviews regarding barriers to adherence were conducted with all CHANTZ participants. Interviews were transcribed and coded for themes by 2 investigators (DRB, GLB) trained in qualitative research, and consensus was reached regarding emerging themes.

**Figure 1.**
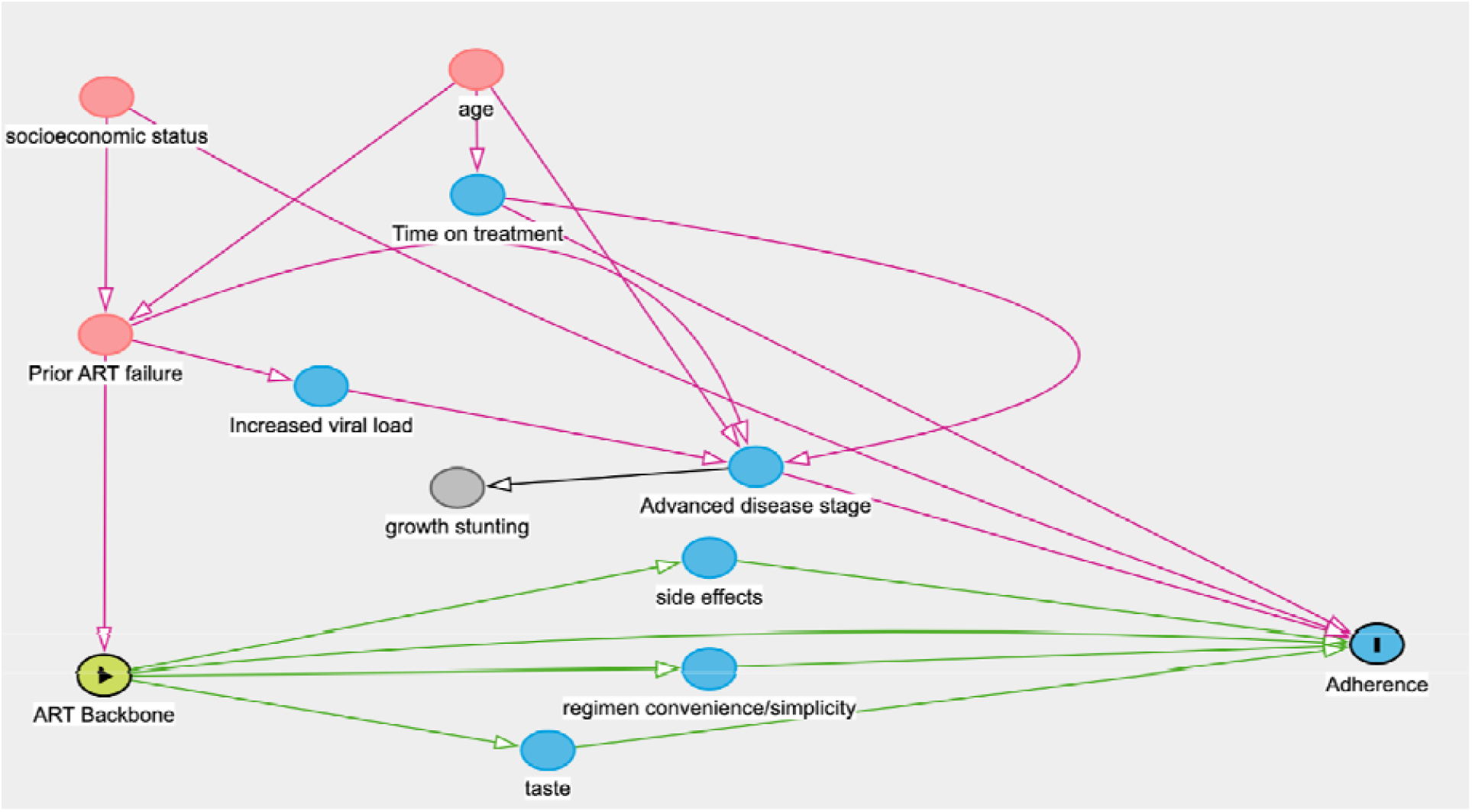
Directed Acyclic Graph showing purported relationship between antiretroviral regimens and medication adherence

## Results

### Demographics & Baseline Group Differences

At baseline, there were 142 children taking Efavirenz, 74 children taking a boosted PI (lopinavir/ritonavir in almost all participants), and 81 children taking an II (in a fixed-dose combination including tenofovir, emtricitabine, and dolutegravir), for a total of 297 children. 3 children were excluded from further analysis due to missing data on their ART regimens. Because HANDZ enrollment took place over five years, there was variable follow-up time (range: 12 - 66 months). Participants taking EFV or a PI at baseline were demographically very similar, while patients taking an II at baseline were younger, had started ART earlier, and had less advanced worst recorded WHO stage (see Table 1). While no differences in self-reported adherence between groups were noted at baseline, participants taking a PI were more likely to have a detectable viral load at baseline (33% of participants vs.16%, p = 0.03).

**Table 1:** Characteristics of Participants on Each ART Regimen at Baseline.

|  |  |  |  |  |
| --- | --- | --- | --- | --- |
| ART | Efavirenz | Protease Inhibitor<br>(Lopinavir/Ritonavir) | Integrase Inhibitor | P-value |
| Total n=297 | n=142 | n=74 | n=81 |  |
| **Mean age at baseline, (SD)** | **11.7 (2.4)** | **11.5 (2.5)** | **10.6 (2.3)** | **0.007** |
| Male sex, n (%) | 77 (54.2%) | 36 (48.6%) | 38 (46.9%) | 0.52 |
| History of severe malnutrition, n (%) | 25 (71.4%) | 20 (66.7%) | 8 (53.3%) | 0.46 |
| **Worst recorded WHO Stage: 1, n (%)** | **19 (13.9%)** | **14 (19.4%)** | **52 (74.3%)** | **<0.001** |
| Worst recorded WHO stage: 2, n (%) | 21 (15.3%) | 7 (9.7%) | 4 (5.7%) |  |
| Worst recorded WHO WHO stage: 3, n (%) | 40 (29.2%) | 14 (19.4%) | 7 (10.0%) |  |
| Worst recorded WHO stage: 4, n (%) | 55 (40.1%) | 37 (51.4%) | 3 (4.3%) |  |
| Current CD4 Count, cells/mm<sup>3</sup> (SD) | 758.9 (330.4) | 820.4 (436.2) | 864.6 (494.3) | 0.18 |
| Lowest recorded CD4 count, cells/mm<sup>3</sup> (SD) | 608.9 (300.6) | 544.0 (356.7) | 681.2 (447.3) | 0.11 |
| **Lowest recorded CD4%, % (SD)** | **26.0 (10.6)** | **21.3 (11.5)** | **24.9 (9.8)** | **0.02** |
| **History of opportunistic infection, n (%)** | **9 (6.4%)** | **6 (8.1%)** | **0 (0.0%)** | **0.04** |
| **Viral load >50 copies/ml (%)** | **13 (9.2%)** | **30 (40.5%)** | **26 (32.1%)** | **<0.001** |
| On first-line therapy at timepoint 1 % | 68% | 70% | 67% | 0.88 |
| **Mean age at initiation of ART years (SD)** | **5.4 (3.7)** | **4.0 (3.1)** | **3.9 (2.5)** | **0.002** |
| History of adherence problems at timepoint 1 | 5% | 7% | 4% | 0.68 |

SD=standard deviation

**Table 2:** Demographics and Clinical Characteristics of Children in the CHANTZ Substudy Taking Protease Inhibitors vs. Other Regimens at Timepoint 2^1^.

| Variable | On II or Efavirenz<br>(n=77) | On Protease Inhibitor<br>(n=18) | P-value |
| --- | --- | --- | --- |
| **Demographics** |  |  |  |
| Mean age, years (IQR) | 14 (12-16) | 14 (12-16) | 0.71 |
| % Female | 42% | 51% | 0.15 |
| **Clinical Characteristics** |  |  |  |
| Most recent CD4 count, cells/mm<sup>3</sup> (SD) | 774 (348) | 821(361) | 0.53 |
| Nadir CD4 count, cells/mm<sup>3</sup> (SD) | 656 (347) | 516 (365) | 0.08 |
| **Adherence & Health History** |  |  |  |
| History of WHO Stage 4 Disease, n (%) | 21 (42%) | 23 (53%) | 0.3 |
| History of Growth Stunting,* n (%) | 10 (20%) | 17 (40%) | 0.01 |
| Most recent viral load undetectable,* n (%) | 47 (92%) | 29 (67%) | 0.002 |
| Most recent viral load, where detectable,* cells/mm<sup>3</sup> (SD) | 951 (2771) | 13,389 (22412) | 0.046 |
| Adherence Problems Noted,* n (%) | 5 (10%) | 12 (28%) | 0.03 |
| Number of days with missed doses in last 7 days,* n (%) | 0 (range: 0) | 0.7 (range: 0-7) | 0.02 |

<sup>1</sup> Age, HIV-related disease severity at baseline, and prior ART failure treated as potential confounders
IQR=inter quartile range
SD=standard deviation
\* indicates statistically significant p-value

### Longitudinal Analysis of Adherence

Children on PI-based regimens were significantly more likely to report adherence problems over the course of follow-up at timepoint 2 (in 28% vs. 10%, p = 0.03) compared to children taking IIs, and noted significantly more days with missed doses in the week prior to interview (median 1 vs. 0, p = 0.02). No significant differences were noted between children taking Efavirenz-based regimens and children taking II-based regimens. Switching from a PI-to an II-based regimen was associated with an increase in median adherence (from 72% of doses taken to 92%, p = 0.01) and an increased proportion of participants having an undetectable viral load at next measurement (from 67% to 78%, p = 0.03). These effects persisted in a linear mixed effects model controlling for age, sex, HIV-related disease severity, and baseline adherence. There was no significant difference in adherence or viral load when switching from Efavirenz to an II.

### Side Effects and Determinants of Adherence

At the time of interview for the CHANTZ substudy (timepoint 2), the majority of patients had already been switched to an II. Thus, CHANTZ included only 18 participants on a PI, 74 participants taking an II, and 3 participants taking Efavirenz. Because of the small number of participants taking Efavirenz, the groups were evaluated as those on PI-based regimens vs. those not on PI-based regimens. The PI group was more likely to report bad medication taste, although this did not reach statistical significance (69% vs. 50%, p = 0.2), and that taste was a problem affecting adherence (22% vs. 4%, p = 0.05). When participants were asked to rate the taste of their medications on a 10-point scale from best (1) to worst (10), patients on protease inhibitors reported significantly worse taste (5.2 vs. 2.5, p = 0.001). In evaluation of side effects, headache was more common in the PI group (17% vs. 4%, p = 0.045), but other side effects were not significantly different between groups.

### Qualitative Analysis

In qualitative interviews, the most common themes given for poor adherence included (1) bad taste of medications, (2) being away from home at the time medications were due, (3) perceived side effects of medication and (4) pill burden/inconvenient scheduling of medications. Many participants reported an unpleasant bitter taste with PIs but described IIs as relatively tasteless.

### Ethics Statement

This study was approved by the institutional review boards of the University of Rochester (protocol #00068985), the University of Zambia (reference #004–08-17), and the National Health Research Association of Zambia. Verbal and written informed consent was required from the parents of all participants to enroll in the study; verbal and written assent was required for all participants 12 and older. Procedures followed were in accordance with the Helinski Declaration of 1975, as revised in 2000.

## Discussion

In this study, we evaluated the effects of a large-scale change from NRTI- and PI-based regimens to II-based regimens in a cohort of children and adolescents in Lusaka, Zambia. We found an association between switching from PI-based regimens to II-based regimens and improved adherence and viral load, as well as an improvement in reported side effects. Self-reported adverse events that led to a decrease in adherence in this cohort included taste disturbances, pill burden, and inconvenient scheduling for taking medications. Pill burden arises from PI-based regimens requiring twice the daily dosage of II-based regimens. Contrary to our original hypothesis, switching from EFV-based regimens to II-based regimens was not associated with any change in adherence, viral load, or reported side effects. Previous studies have characterized the common side effects experienced in by individuals on PI-based regimens, as well as in individuals on NNRTI- or II-based regimens. In prior studies, patients taking PI regimens have noted adverse events such as nausea, vomiting, headaches, and diarrhea among other adverse events [1,2,15]. Similar adverse events have been reported with NNRTI- and integrase inhibitor-based regimens, albeit at lower rates. Protease inhibitors have higher rates of reported adverse events than other antiretrovirals, although some studies have suggested that poor adherence to protease inhibitors may be less likely to result in treatment failure in the course of the disease than poor adherence to other regimens [7, 19, 21].

To our knowledge, this is one of the first studies that has evaluated the effect on adherence of switching children and adolescents from PI- and NNRTI-based regimens to II-based regimens. This study should provide some reassurance regarding the large-scale transition to IIs taking place in Sub-Saharan Africa. In the future, more effective adherence interventions to increase adherence in adolescents are crucial, as this population represents one of the major groups at risk for treatment failure. These interventions must target different social factors impacting adherence such as depression, family stressors, and poverty, to allow for sustainable and scalable change in community health. In addition, medication taste likely contributes to poor adherence and viral escape in this age group, so palatability should be a priority in drug development for pediatric HIV.

This study had a number of limitations. We were only able to evaluate the effect of switching to IIs; switching from IIs to other regimens was not observed due to very low rates of treatment failure on IIs. The effect of switching the ART regimen itself could potentially cause a temporary increase in adherence unrelated to the particular regimen. However, we consider this unlikely as this effect was observed only in participants switching from PIs. Another limitation of this study includes the relatively short follow-up period after switching to IIs. Continued surveillance of this population as part of the HANDZ study will allow for a more comprehensive picture of the adherence rates and factors influencing adherence within this patient cohort. In addition, almost all participants on PIs were taking lopinavir/ritonavir, and other PIs may not have the same issues. Finally, this study was conducted at a single urban referral center, and results may not generalize to populations with substantially different demographics.

In conclusion, switching children and adolescents from PI-based regimens to II-based regimens has a number of advantages, including improved adherence, less burdensome side effects, and more optimal viral suppression. It is important to note that taste of medication is likely an important factor affecting adherence and viral suppression in CAWH. Drug development for pediatric HIV should include palatability of medication as an important consideration.

## Data Availability

All data produced in the present study are available upon reasonable request to the authors.

## Acknowledgments

Research reported in this publication was supported by the National Institute Of Mental Health of the National Institutes of Health under Award Number R21MH122238 and K23NS117310. The content is solely the responsibility of the authors and does not necessarily represent the official views of the National Institutes of Health.

## Data Availability Statement

The data used in this study are available upon reasonable request from David R. Bearden.

## Notes

**Conflicts of Interest and Source of Funding:** Research reported in this publication was supported by the National Institute Of Mental Health of the National Institutes of Health under Award Number R21MH122238 and K23NS117310. The content is solely the responsibility of the authors and does not necessarily represent the official views of the National Institutes of Health. The authors have no conflicts of interest to disclose.

### Competing Interest Statement

The authors have declared no competing interest.

### Author Declarations

The institutional review boards of the University of Rochester (protocol #00068985), the University of Zambia (reference #004 08 17), and the National Health Research Association of Zambia gave ethical approval for this work.

